# Krebs von den Lungen 6 levels in COVID-19 ICU Patients are Associated with Mortality

**DOI:** 10.1101/2021.11.17.21266464

**Authors:** Giuliana Scarpati, Daniela Baldassarre, Graziella Lacava, Filomena Oliva, Gabriele Pascale, Massimo Boffardi, Pasquale Pagliano, Vincenzo Calabrese, Giovanni L. Tripepi, Ornella Piazza

## Abstract

**Rationale:** Krebs von den Lungen 6 (KL-6) is a high molecular weight mucin-like glycoprotein produced by type II pneumocytes and bronchial epithelial cells. Elevated circulating levels of KL-6 may denote disorder of the alveolar epithelial lining.

**Objective:** Aim of this study was to verify if KL-6 values may help to risk stratify and triage severe COVID-19 patients.

**Methods:** We performed a retrospective prognostic study on 110 COVID-19 ICU patients, evaluating the predictive role of KL-6 for mortality.

**Measurements and Main Results:** The study sample was divided in two groups related according to the median KL-6 value [Group A (KL-6 lower than the log-transformed median (6.73)) and Group B (KL-6 higher than the log-transformed median)]. In both linear and logistic multivariate analyses, ratio of arterial partial pressure of oxygen to fraction of inspired oxygen (P/F) was significantly and inversely related to KL-6. Death rate was higher in group B than in group A (80.3 versus 45.9%) (p<0.001), Accordingly, the Cox regression analysis showed a significant prognostic role of KL-6 on mortality in the whole sample as well as in the subgroup with SOFA lower than its median value.

**Conclusions:** At ICU admission, KL-6 serum level was significantly lower in the survivors group. Our findings shown that, in severe COVID19 patients, elevated KL-6 was strongly associated with mortality in ICU.

## Introduction

COVID-19 mainly affects the respiratory system by causing a low oxygenation index in about 14% of patients (1). Even if COVID-19 clinical manifestations are mild in some patients, with no complaint of dyspnea, no significant increase in respiratory rate, and no respiratory distress (the so called “happy hypoxia”) (2), other patients (about 5%, as reported by Wu et Al (1) in a study including 72314 persons with COVID-19 in China) rapidly progress to respiratory failure and acute respiratory distress syndrome (ARDS). COVID-19 ARDS patients should be admitted to the intensive care unit (ICU), the elderly and those with comorbidities being at highest risk of death.

ARDS, as defined by Berlin criteria (3), is a complex syndrome, caused by very different diseases, and therefore it is not surprising that COVID-19-related ARDS patients present some differences with ARDS caused by other factors (4).

ARDS has been also classified among the Interstitial Lung Diseases (ILDs), among which are listed, i.e., the collagen vascular disease-associated interstitial pneumonia and the hypersensitivity pneumonia (5). Interstitial lung diseases (ILDs) are a group of pulmonary disorders characterized by diverse patterns of inflammation and fibrosis in the interstitium of the lung. ILDs express a shared pathophysiological change, that is the type II pneumocytes damage or transformation. In ARDS patients, the disorder of the alveolar epithelium has been described and it is known that lung epithelial intactness and functionality are important elements for their clinical consequences, since when alveolar type II cells are damaged, removal of alveolar edema fluid is compromised. Also, injury to type II cells reduces the production and turnover of surfactant, which is associated to poor outcome (6).

The Krebs von den Lungen 6 (KL-6) protein is a high molecular weight mucin-like glycoprotein, produced by type II pneumocytes and bronchial epithelial cells.

Since elevated circulating levels of KL-6 indicate disruption of the alveolar epithelial lining, we hypothesized that it could be advantageous analyzing KL-6 in a group of severe COVID-19 positive patients, to better risk stratify and triage them.

Central question of this study was investigating the association between KL6 serum levels, clinical severity scores and ICU COVID-19 mortality.

## Methods

This is a single-center, retrospective observational study performed at Salerno University COVID Hospital “G. Da Procida”, which was designated as a COVID-only medical center in Salerno, Italy, on the 8^th^ of October 2020.

The ICU admission was reserved to patients defined critically ill. According to National Institute of Health (NIH) guidelines (7), critical patients are identified as:

- *Severe Illness:* Individuals who have SpO2 <94% on room air at sea level, a ratio of arterial partial pressure of oxygen to fraction of inspired oxygen (P/F) <300 mm Hg, respiratory frequency >30 breaths/min, or lung infiltrates >50%.
- *Critical Illness:* Individuals who have respiratory failure, septic shock, and/or multiple organ dysfunction.

150 patients were admitted during the II pandemic wave in the Covid ICU from 8^th^ of October 2020 to 8^th^ of June 2021, mean age was 67.40 ± 11.18 year. 110 subjects were males (73%), 91 were non-survivors, representing a 61% hospital mortality rate. Endotracheal intubation was necessary in 45 patients (30%) while noninvasive ventilation (facial mask/helmet NIV) was used in 105 patients (70%). Baseline characteristics of source population are listed in table 1.

**Table 1:** Baseline characteristics of source population. Data are presented as mean ± SD for continuous variables normally distributed, percentage are reported for categorical variables. NIV: non-invasive ventilation; IOT: orotracheal intubation.

|  | **Survivors (n = 59)** | **Non - Survivors (n = 91)** |
| --- | --- | --- |
| Gender (M/W) (%) | 79/21 | 69/31 |
| Age (years) | 63.38 ± 12.4 | 69.9 ± 9.7 |
| Hypertension (%) | 36 | 49 |
| Obesity (%) | 11 | 14 |
| Diabetes (%) | 23 | 29 |
| Cardiovascular pathologies (%) | 13 | 23 |
| Respiratory pathologies (%) | 13 | 20 |
| Autoimmune disease (%) | 0 | 4 |
| Oncological pathologies (%) | 4 | 5 |
| Length of stay (ICU days) | 7 ± 5 | 8 ± 5 |
| White blood cells (n/μL) | 9837.77 ± 4109.87 | 11803.86 ± 7348.52 |
| Procalcitonin (pg/ml) | 0.24 ± 0.51 | 4.26 ± 20.60 |
| SOFA score | 4.19 ± 2.29 | 6.34 ± 2.93 |
| SAPS II | 21.71 ± 5.32 | 30.95 ± 11.23 |
| P/F (PaO<sub>2</sub>/FiO<sub>2</sub>): |  |  |
| <100 (%) | 38 | 73 |
| 100-200 (%) | 55 | 53 |
| >200 (%) | 13 | 5 |
| Respiratory support: |  |  |
| NIV (%) | 66 | 37 |
| IOT (%) | 5 | 45 |

**Table 2.** Baseline KL6 at ICU admission. ARDS was diagnosed at the ICU admission as severe in 49% of cases, moderate in 38%, mild in 4%, while the remaining patients were diagnosticated as pneumothorax/pnemomediastinum or pleural effusions

Tab 2
|  | Non survivors<br>(91 pts) | Survivors<br>(59 pts) | Endotracheal<br>Intubation (44 pts) | NIV (n 106 pts) |
| --- | --- | --- | --- | --- |
| KL-6 U/ml<br>(mean ± ds) | 1516±1236 | 845 ±936 | 1693±1305 | 1081±1076 |
|  | p= 0,0004 |  | p= 0.012 |  |

**Table 3:** Group A (KL-6 lower than the log median) and Group B (KL-6 higher than the median). The two groups differed for P/F value (p=0.001), SOFA (p=0.003). Correlation analysis fully confirmed the results of the categorical approach (see last column).

Tab 3
| Variables | complete<br>sample n=122<br>median values | Group A<br>n=61 | Group B<br>n=61 | p |
| --- | --- | --- | --- | --- |
| Age | 67±11 | 67±13 | 68±9 | 0.443 |
| Sex M/F | 31/91 | 15/46 | 16/45 | 0.907 |
| hypertension | 58(47.5%) | 24 (39.3%) | 34 (55.7%) | 0.102 |
| Obesity | 17(13.9%) | 9 (14.8%) | 8(13.1%) | 0.794 |
| diabetes | 36(29.5%) | 20 (32.8%) | 16(26.2%) | 0.552 |
| CV<br>pathologies | 26(21.3%) | 12 (19.7%) | 14 (23.0%) | 0.825 |
| Respiratory p | 23(18.9%) | 11 (18.0%) | 12 (19.7%) | 0.817 |
| Autoimmune<br>disease | 2(1.6%) | 1 (1.6%) | 1 (1.6%) | 1.000 |
| Malignancies | 6(4.9%) | 3 (4.9%) | 3 (4.9%) | 1.000 |
| P/F | 126±59 | 146±69 | 108±40 | **0.001** |
| SOFA | 5[3-7] | 4 [3-6] | 6[4-8] | **0.003** |
| WBC day1<br>10<sup>3</sup>/mm<sup>3</sup> | 9.8[7.4-13.2] | 9.2 [7-11.8] | 11.5 [8.1-14.3] | 0.169 |
| PCT ng/ml | 0.15[0.08-0.71] | 0.11[0.07-0.32] | 0.32 [0.1-1.0] | 0.088 |

**Table 4:** Univariate linear and logistic regression analysis. The two groups differed for P/F value (p=0.003) and SOFA (p=0.005).

Tab 4
| Variables | Linear regression |  |  | Logistic Regression |  |  |
| --- | --- | --- | --- | --- | --- | --- |
| | $\beta$ | 95% CI | p | Odds ratio | 95% CI | p |
| Age | 0.001 | -0.01/0.02 | 0.916 | 1.01 | 0.98/1.05 | 0.424 |
| Sex M/F | 0.101 | -0.255/0.576 | 0.576 | 0.92 | 0.41/2.07 | 0.917 |
| hypertension | 0.206 | -0.103/0.514 | 0.190 | 1.94 | 0.94/3.99 | 0.071 |
| Obesity | 0.033 | -0.415/0.481 | 0.885 | 0.87 | 0.31/2.43 | 0.872 |
| diabetes | -0.002 | -0.342/0.339 | 0.992 | 0.73 | 0.33/1.59 | 0.428 |
| CV pathologies | 0.183 | -0.194/0.561 | 0.338 | 1.21 | 0.51/2.90 | 0.659 |
| Respiratory p | 0.137 | -0.259/0.533 | 0.495 | 1.11 | 0.45/2.76 | 0.817 |
| Autoimmune disease | -0.165 | -1.38/1.057 | 0.789 | 1.00 | 0.06/16.4 | 1.00 |
| Malignancies | -0.083 | -0.801/0.635 | 0.819 | 1.00 | 0.194/5.161 | 1.00 |
| P/F | -0.004 | -0.006/-0.001 | 0.007 | 0.99 | 0.979/0.995 | **0.002** |
| SOFA | 0.071 | 0.021/0.122 | 0.006 | 1.21 | 1.06/1.39 | **0.005** |
| WBC day1<br>10<sup>3</sup>/mm<sup>3</sup> | 0.026 | 0.001/0.05 | 0.038 | 1.06 | 0.99/1.13 | 0.065 |
| PCT ng/ml | 0.00023 | -0.010/0.011 | 0.925 | 1.012 | 0.98/1.05 | 0.483 |

**Table 5:** In both linear and logistic multivariate analyses, P/F was significantly and inversely related to KL-6

**Tab 5**
| Variables | Linear regression |  |  | Logistic regression |  |  |
| --- | --- | --- | --- | --- | --- | --- |
| | $\beta$ | 95% CI | p | OR | 95% CI | p |
| Age | -0.003 | -0.018/0.012 | 0.695 | 1.004 | 0.96/1.04 | 0.834 |
| Sex M/F | 0.153 | -0.218/0.525 | 0.415 | 1.18 | 0.46/3.03 | 0.740 |
| P/F | -0.003 | -0.006/-0.0005 | 0.023 | 0.99 | 0.978/0.998 | **0.009** |
| SOFA | 0.041 | -0.016/0.098 | 0.158 | 1.14 | 0.97/1.32 | 0.114 |
| WBC day1<br>10<sup>3</sup>/mm<sup>3</sup> | 0.020 | 0.004/0.044 | 0.107 | 1.04 | 0.97/1.12 | 0.250 |

122 patients with KL-6 measurement were included in our study, 28 patients were excluded because KL-6 values at ICU admission were not available. The non-survivors (n=77) were older (70 ± 10 yrs) than the survivors (63 ± 13 yrs).

KL-6 was measured with commercial kit produced by Tosoh (ST AIA-PACK KL6) following producer instructions.

Campania SUD Institutional Review Board approved this study (protocol ID 0008402010) as minimal risk research.

Numerous variables were analyzed, including age, sex, body mass index, comorbidities and severity scores (P/F ratio, Sequential Organ Failure Assessment (SOFA) score (8)), KL-6, procalcitonin, white blood cells count. These biomarkers were measured at the time of critical care admission. The primary outcome was ICU mortality.

## Statistical Analysis

The distribution of variables was evaluated by Kolgomorov-Smirnov test and with graphical evaluation. Variable with positively skewed distribution were log-transformed. At baseline, continuous variables were compared by student T-test or Mann-Whitney test, as appropriate, and dummy variables by Pearson’s Chi-Square. Linear regression or logistic regression analyses were performed to assess the association between KL-6 and other variables. Kaplan-Meier analysis and Cox regression were performed in the whole sample as well as patients with KL6 below and above the median. We used SPSS version 24, Chicago, Illinois, USA.

## Results

At ICU admission, KL-6 serum level was significantly lower in the survivors group (median value 545 U/ml vs 1070 U/ml in those patients who did not survive at 28 day after ICU admission) (fig 1) and in patients that were managed with NIV for the whole lenght of ICU stay (711 U/ml vs 1073 U/ml in patients who required endotracheal intubation at the admission or during the ICU stay) (tab 2). KL-6 was not correlated with gender and there was no difference in median values at the admission (median males 942.5 U/ml median females 840 U/ml), but in our population women represented the 25% of the population. SOFA score at ICU admission was (5.5±3.0); a SOFA score >3 was associated to poor prognosis (sensitivity 56,6 %; specificity 87,5 %) (Fig 2).

**Fig 1.**
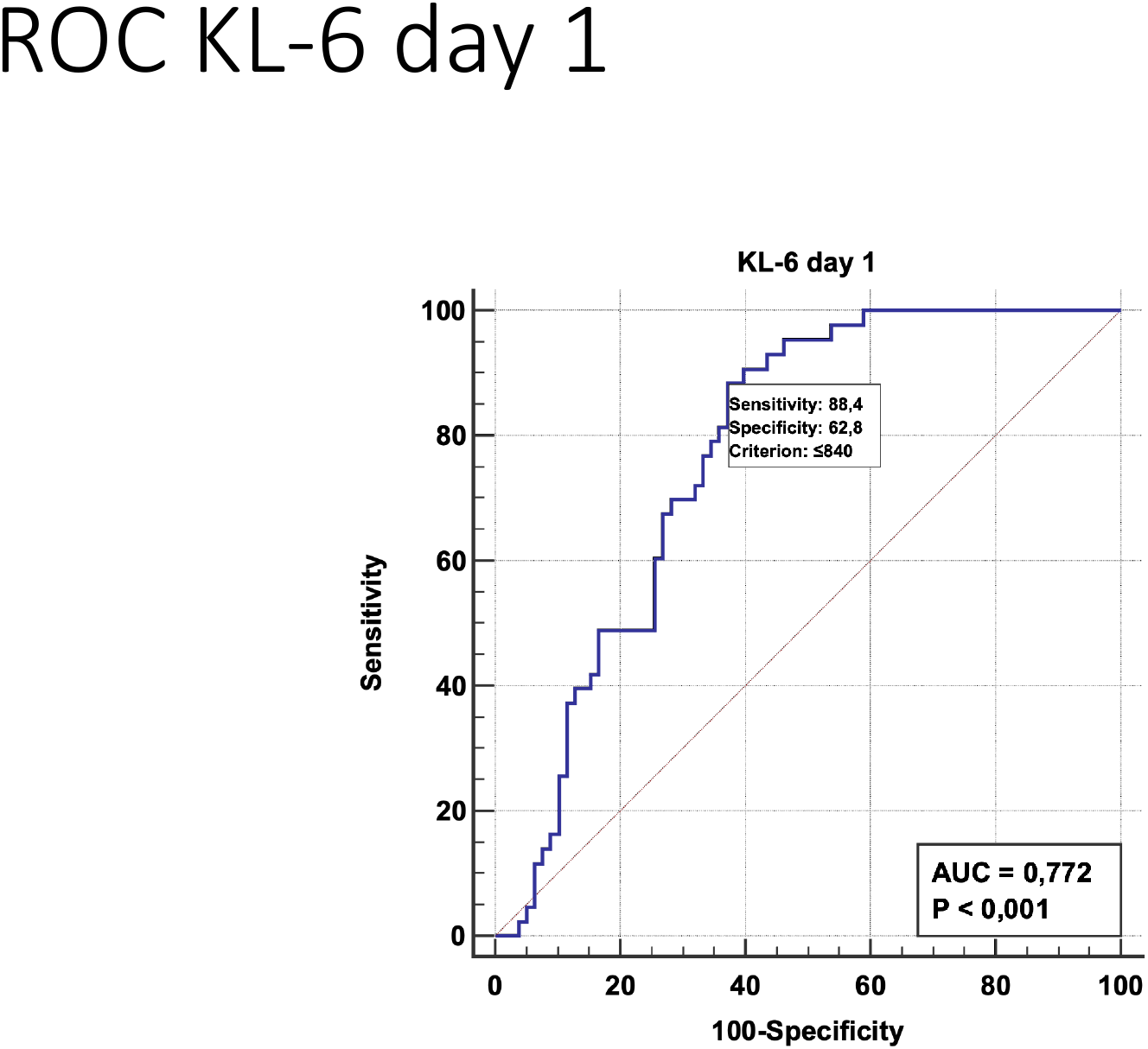
Dot plot diagram comparing KL-6 values measured on admission comparing patient who survived with patient who did not survive.

**Fig 2:**
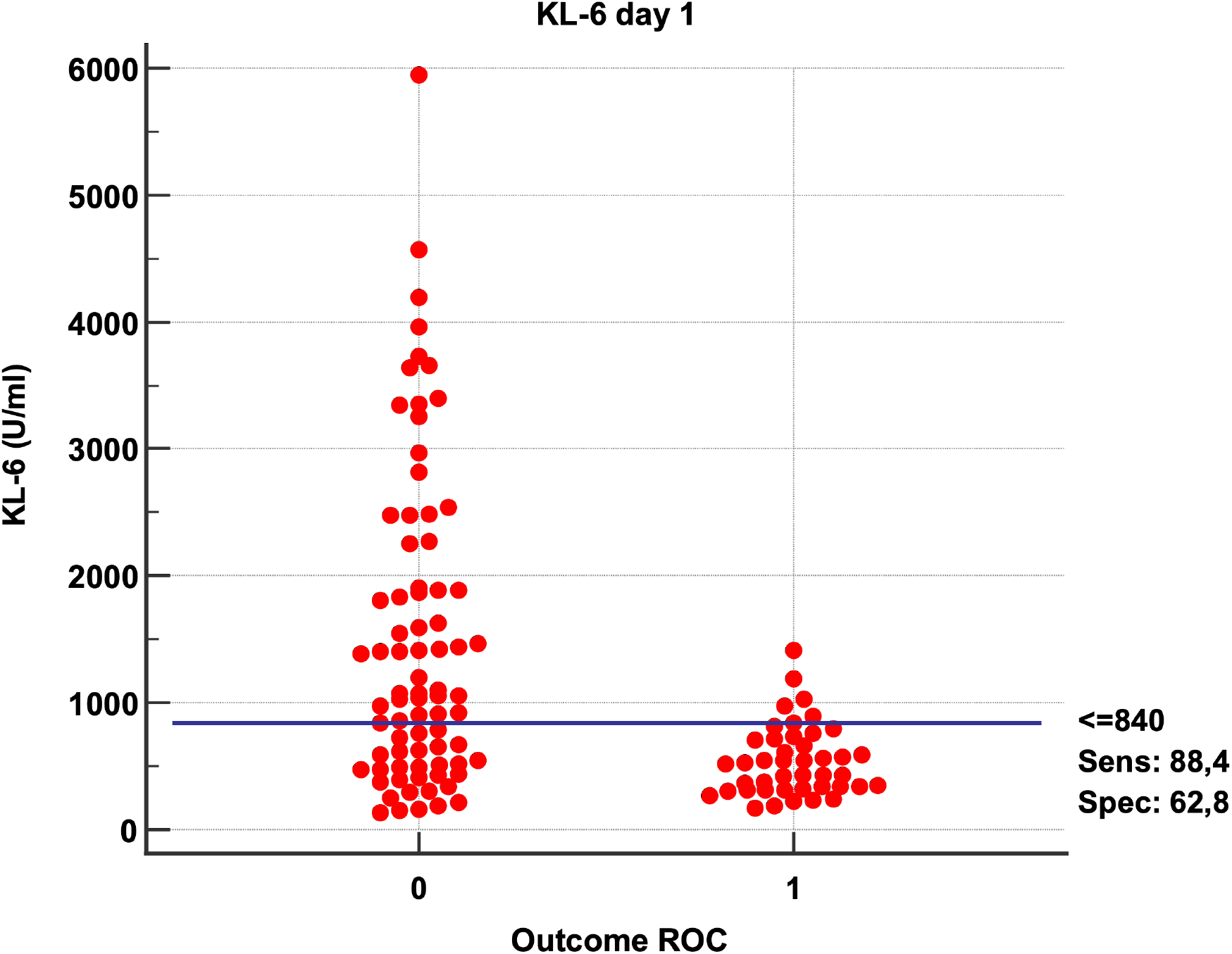
ROC curve analysis of SOFA score on the day of ICU admission. Patients with a SOFA score >3 have a poor prognosis (sensitivity 56,6 %; specificity 87,5 %).

This restricted study sample (122 pts) was divided in two groups related according to the median log-transformed KL-6 value (6.73 U/ml) [Group A (KL6 lower than the median) and Group B (KL6 higher than the median)]: mortality rate was higher in group B than in group A (80.3 versus 45.9%) (p<0.001). Baseline features are summarized in Tab. 3, univariate linear and logistic regression analysis provided similar results (Tab 4). In both linear and logistic multivariate analyses, only P/F was significantly and inversely related to KL-6. (Tab 5).

Kaplan-Meier analysis showed a significant higher mortality in patients who had a KL-6 value at admission > 6.73 U/ml (Group B), both in the whole study sample as well as in those patients who had high KL-6 but SOFA score < 5.0 (median value of SOFA in this population). Conversely, in the subgroup with SOFA > 5.0 (higher than the SOFA median), this impact of KL-6 on mortality was not evident (Fig 3). Accordingly, the Cox regression analysis showed a significant prognostic role of KL-6 on mortality in the whole sample (HR:1.78, 95% CI 1.12-2.82, p=0.02) as well as in the subgroup with SOFA lower than the corresponding median value (HR:1.86, 95%CI 1.02-3.44, p=0.045), but not in the subgroup of patients with SOFA above the median (HR 0.96, 95%CI 0.69-1.34, p=0.96). Therefore, SOFA acted as a significant effect modifier of the KL-6/mortality link (P=0.048).

**Fig. 3.**
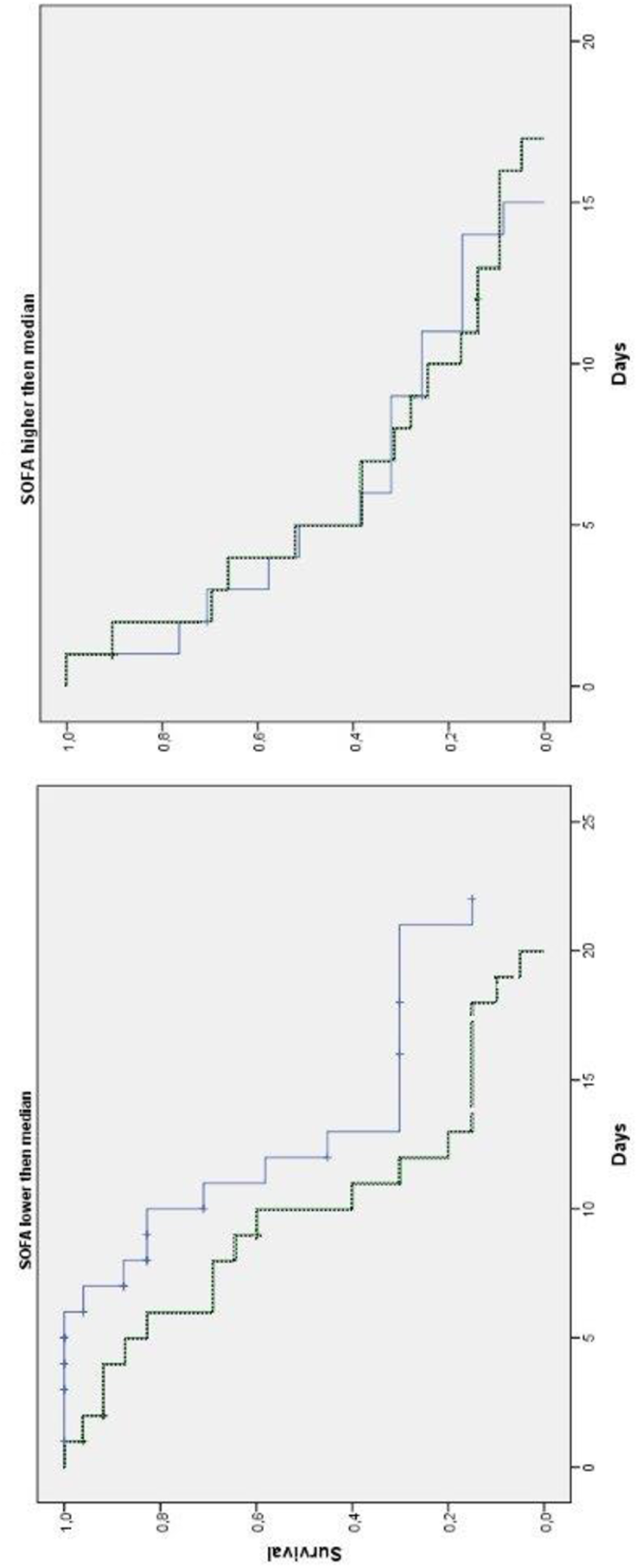
Kaplan-Meier curves in two subgroups splitted from SOFA median (SOFA median value=5): lower than median (3a) and SOFA value higher than median (3b). Dashed blue line represents Group A; the solid green line represents group B. Fig 3a (SOFA lower than 5) reported a higher cumulative probability of survival on group A (median 12 days) as compared to group B (median 10 days). Conversely, 3b imagine (SOFA higher than 5) reported a similar cumulative probability of survival was similar in both groups (median 6 vs 5 days in group A and group B, respectively). KL-6 was a negative prognostic marker in the subgroup with SOFA lower than 5, while when SOFA was higher than 5, in the most severe patients, this prognostic power was not statistically significant.

## Conclusions

This retrospective prognostic study correlated clinical characteristics and KL-6 values with disease severity in COVID-19 patients receiving critical care consultations during the second wave of the pandemic in a COVID dedicated Hospital.

KL-6 breaches at the S-S bond near the epithelial membrane surface and becomes distributed in pulmonary epithelial lining fluid (ELF). This glycoprotein is mainly expressed on alveolar type II cells in the lung, and it is produced more prominently by proliferating, regenerating, or injured type II cells than by healthy type II cells. The presence of KL-6 has been used to monitor severity of disease in idiopathic pulmonary fibrosis (6). Our findings revealed that elevated KL6 was most strongly associated with mortality in ICU, thereby contributing to the growing body of evidence for the utility of KL6 in the context of COVID-19 infection (9)

Unfortunately, this tragic second wave found us, as Salerno University Hospital, in October 2020 not yet ready for the fight against COVID-19 and a temporary 8 bed ICU was arranged, as soon as possible, in a general ward in disuse (10). We attribute the high mortality in our patient population to their disease severity, advanced age and the limitation in resources available, given the huge and coincident influx of critically ill patients (11). It is important to note that the south of Italy community has a high prevalence of severe COVID-19 risk factors like diabetes, hypertension and obesity (12). The presence of these cofactors could account for the high mortality in this patient population.

Over time, ICU mortality has lessened world-wide and this may be due to new treatments (13) like steroids (14) and Tociluzimab (15).

Our study aimed to identify factors that the intensivist could use at the time of consult to risk stratify COVID-19 patients. The use of biomarkers to predict disease severity has proven essential for resource allocation, particularly for respiratory support needs.

The main limitation of this research is that is an observational study with retrospective enrolment. For this reason, our analysis was performed including only patients with complete anamnestic and laboratory data; moreover, we think that the KL-6 value as prognostic index should be tested in different population, i.e. at Emergency Room admission.

## Data Availability

All data produced in the present study are available upon reasonable request to the authors

## References

1. Wu Z MJ. Characteristics of and important lessons from the coronavirus disease 2019 (COVID-19) outbreak in China: summary of a report of 72,314 cases from the Chinese Center for Disease Control and Prevention. JAMA 2020;; 323: 1239–1242.

2. Tobin MJ LF, Jubran A.. Why COVID-19 silent hypoxemia is baffling to physicians. Am J Respir Crit Care Med 2020; 202: 356–360.

3. Definition Task Force ARDS RV, Rubenfeld GD, et al. Acute respiratory distress syndrome: the Berlin definition.. JAMA 2012; 307: 2526–2533.

4. Li X MX. Acute respiratory failure in COVID-19: is it “typical” ARDS?. Crit Care 2020; 24:.

5. Ishikawa N HN, Yokoyama A, Kohno N. Utility of KL-6/MUC1 in the clinical management of interstitial lung diseases.. Respir Investig 2012; 50: 3–13.

6. Ishizaka A, Matsuda, T., Albertine, K. H., Koh, H., Tasaka, S., Hasegawa, N., Hashimoto, S. Elevation of KL-6, a lung epithelial cell marker, in plasma and epithelial lining fluid in acute respiratory distress syndrome.. American Journal of Physiology - Lung Cellular and Molecular Physiology 2004; 286: L1088–L1094.

7. National Institute of Health. Coronavirus Disease 2019 (COVID-19) Treatment Guidelines.

8. Vincent JL, Moreno R, Takala J, Willatts S, De Mendonca A, Bruining H, Reinhart CK, Suter PM, Thijs LG. The SOFA (Sepsis-related Organ Failure Assessment) score to describe organ dysfunction/failure. On behalf of the Working Group on Sepsis-Related Problems of the European Society of Intensive Care Medicine. Intensive Care Med 1996; 22: 707–710.

9. d’Alessandro M, Bergantini L, Cameli P, Curatola G, Remediani L, Bennett D, Bianchi F, Perillo F, Volterrani L, Mazzei MA, Bargagli E, Siena C. Serial KL-6 measurements in COVID-19 patients. Intern Emerg Med 2021; 16: 1541–1545.

10. Mojoli F, Cutti, S., Mongodi, S. The potential role of ICU capacity strain in COVID-19 mortality: comparison between first and second waves in Pavia, Italy. J Anesth Analg Crit Care 2021; 1: 8.

11. Rubinson L. “Intensive care unit strain and mortality risk among critically ill patients with covid-19-there is no “me” in covid.” JAMA Netw Open 2021.

12. Mancusi C, Grassi, G., Borghi, C. et al. Clinical Characteristics and Outcomes of Patients with COVID-19 Infection: The Results of the SARS-RAS Study of the Italian Society of Hypertension. High Blood Press Cardiovasc Prev 2021; 28: 5–11.

13. Pagliano P SG, Sellitto C, Conti V, Spera AM, Ascione T, Piazza O, Filippelli A.. Experimental Pharmacotherapy for COVID-19: The Latest Advances.. J Exp Pharmacol 2021; 13: 1–13.

14. Tomazini BM, I. S. Maia, A. B. Cavalcanti, O. Berwanger, R. G. Rosa, V. C. Veiga, A. Avezum, R. D. Lopes, F. R. Bueno, M. Silva, et al. Effect of dexamethasone on days alive and ventilator-free in patients with moderate or severe acute respiratory distress syndrome and covid-19: The codex randomized clinical trial. JAMA Netw Open 2020; 324: 1307–1316.

15. Hermine O, X. Mariette, P. L. Tharaux, M. Resche-Rigon, R. Porcher, P. Ravaud and C.-C. Group. Effect of tocilizumab vs usual care in adults hospitalized with covid-19 and moderate or severe pneumonia: A randomized clinical trial. JAMA Intern Med 2021; 181: 32–40..

